# Prevalence of non-tuberculous mycobacteria among people presumed to have tuberculosis, positive for acid-fast bacilli in Mali

**DOI:** 10.1101/2024.04.16.24304822

**Authors:** Aissata Boubakar Cisse, Anna S. Dean, Armand Van Deun, Jelle Keysers, Willem-Bram De Rijke, Mourad Gumusboga, Hawa Samake, Seydou Arama, Bassirou Diarra, Ibrahim Djilla, Fatoumata N. Coulibaly, Hawa Simpara, Mamadou Berthe, Khadidia Ouattara, Yacouba Toloba, Ibrehima Guindo, Bouke de Jong, Leen Rigouts

**Author notes:** Deceased in September 2023 (Armand Van Deun). Corresponding author (ABC).

## Abstract

**Background:** Non-tuberculosis mycobacteria (NTMs) are environmental agents that can cause opportunistic pulmonary disease in humans and animals which is often misdiagnosed as tuberculosis (TB). In this study, we describe the cases of NTMs identified during the first national anti-TB drug-resistance survey conducted in Mali, and associated risk factors.

**Methods:** Sputum was collected from people presenting for pulmonary TB diagnosis, from April to December 2019, regardless of age. Microscopy-positive patients were enrolled and were tested by GeneXpert MTB/RIF. Cases that tested negative for the *Mycobacterium tuberculosis* complex (MTBc) were tested for presence of mycobacteria by amplification of the IS*6110* and 16SrRNA genes through double quantitative real-time PCR, followed by nested PCR and Sanger sequencing of the IS*6110*-negative samples for NTM species identification.

**Results:** A total of 1,418 sputum smear-positive patients were enrolled, including 1,199 new cases, 211 previously treated cases and 8 whose previous treatment history was unknown. Based on the results of GeneXpert MTB/RIF and in-house PCR methods, 1331 (93.9%) patients were positive for MTBc, 48 (3.4%) for NTMs and for 39 (2.7%) no species identification was possible. Advanced age (65 and over) (OR 8.8, p=0.001) and previous TB treatment (OR 3.4 and p=0.016) were the risk factors statistically associated with NTM detection. *M. avium complex* (MAC) was the predominant NTM species, detected in 20 cases.

**Conclusion:** Detection of NTMs in people presumed to have TB is an ongoing challenge, confounding correct TB diagnosis. Concomitant use of microscopy and GeneXpert testing among at-risk individuals could lessen confusion.

## 1. Introduction

The *Mycobacterium* genus groups together species of the *Mycobacterium tuberculosis* complex (MTBc) responsible for tuberculosis (TB) in humans and animals, *M. leprae*, the causative agent of leprosy, and finally, the species of mycobacteria commonly known as atypical or non-*tuberculosis* or mycobacteria (NTM) including *M. ulcerans* responsible for Buruli ulcer. NTMs are recognised as environmental bacteria(1–4), yet some species have been associated with pulmonary diseases with symptoms similar to TB (5). All mycobacteria are acid-fast bacilli (AFB) detectable by microscopy after specific staining, yet an AFB-positive result by sputum smear cannot specify the species. A negative result for the MTBc by GeneXpert MTB/RIF (Xpert hereafter) when performed on AFB-positive sputum can suggest an NTM infection (6). Cases of NTM infection in Mali were described before the introduction of Xpert into the TB diagnostic algorithm, particularly in patients who have been previously treated for TB (7).

Unlike members of the MTBc, NTMs are not obligate parasites of humans. They are normal residents of soil and water. Over 170 species of NTM have been officially recognized(8), of which only 25 species have been strongly associated with disease in humans. Because their habitat exposes humans to these bacteria every day, the diagnosis of NTM disease must be differentiated from simple colonization or contamination of clinical specimens, for example by tap water. Identification to the species level is also important as treatment differs from one NTM species to another.

In this article, we describe the proportion of MTBc and of NTM detected in patients undergoing initial treatment or retreatment for TB in Mali, in the context of a drug-resistance survey. We also looked for risk factors associated with NTM detection.

## 2. Methods

### 2.1. Design of the study

The study was conducted from April to December 2019 in the context of the first national TB drug-resistance survey carried out in Mali (9). Consecutive eligible patients were recruited from 78 of 86 TB diagnostic laboratories in the country, representing 91% of the country’s TB laboratory network. Laboratories that notified less than 10 new bacteriologically confirmed pulmonary TB cases in 2017 were excluded for logistic reasons. Eligible participants were defined as people undergoing evaluation for pulmonary TB, both with and without a history of prior treatment, regardless of age. The sample size of 994 new cases was calculated to meet the primary objective of the survey for measuring the percentage of new pulmonary TB cases resistant to rifampicin, for which a recruitment period of 4 months was estimated to be required(10).

### 2.2. Sample collection and patient enrolment

Two sputum samples, each of approximately 5 mL, were initially collected from each patient in the participating laboratories. Smears were made directly from freshly collected sputum, stained according to the acid-fast staining technique used at the center (auramine for most sites and Ziehl-Neelsen for several small centers) and following international guidelines for diagnosing TB by microscopy(11). In case one of these two sputum samples was AFB positive, the patient was included in the drug-resistance survey after giving informed consent, and an additional sample was collected.

The nurse completed the study questionnaire with the recruited patient. The questionnaire covered socio-demographic data, and information on previous TB treatment and pulmonary disease history (duration and chronicity of respiratory signs).

### 2.3. Initial Xpert testing and specimen storage

The first two sputum samples collected were preserved in 5 mL of Omnigene Sputum decontamination reagent (DNA Genotek Inc, Canada) according to the manufacturer’s recommendations (12), while the third sample was preserved in 10 mL of 96% ethanol. The conditioned samples were transported by public transport at ambient temperature in triple packaging to the National Tuberculosis Reference Laboratory in Bamako.

To detect MTBc, the GeneXpert MTB/RIF assay (Cepheid, CA, USA) was performed at the National Tuberculosis Reference Laboratory, using 2mL of the sample preserved in Omnigene, in accordance with the manufacturer’s recommendations (Cepheid and Genotek).

Leftovers of the samples were then stored in cryotubes in the form of sediment obtained after centrifugation at 4000g for 15 minutes and mixed with 96% alcohol at an equal volume (0.5 mL sediment and 0.5 mL ethanol).

### 2.4. Molecular detection and identification of NTM species

For molecular detection and identification of NTM species, AFB-positive/MTBc-negative samples were transported at ambient temperature to the WHO Supranational Tuberculosis Reference Laboratory at the Institute of Tropical Medicine (ITM) in Antwerp, Belgium. Upon arrival at ITM, ethanol-preserved samples were centrifuged at 4000 g for 15 minutes, digested by overnight proteinase K incubation, followed by lysis using an internal lysis buffer, after which DNA was extracted using the Modified Maxwell-LEV® automated extraction method (Promega, USA) (13)

A duplex quantitative real time polymerase chain reaction (qPCR) was performed, targeting the IS*16110* gene for the detection of the MTBc and the 16S rRNA gene to detect all species of the *Mycobacterium* genus. The primers and probes used are presented in Table 1, while interpretation of results is depicted in Table 2.

**Table 1.**
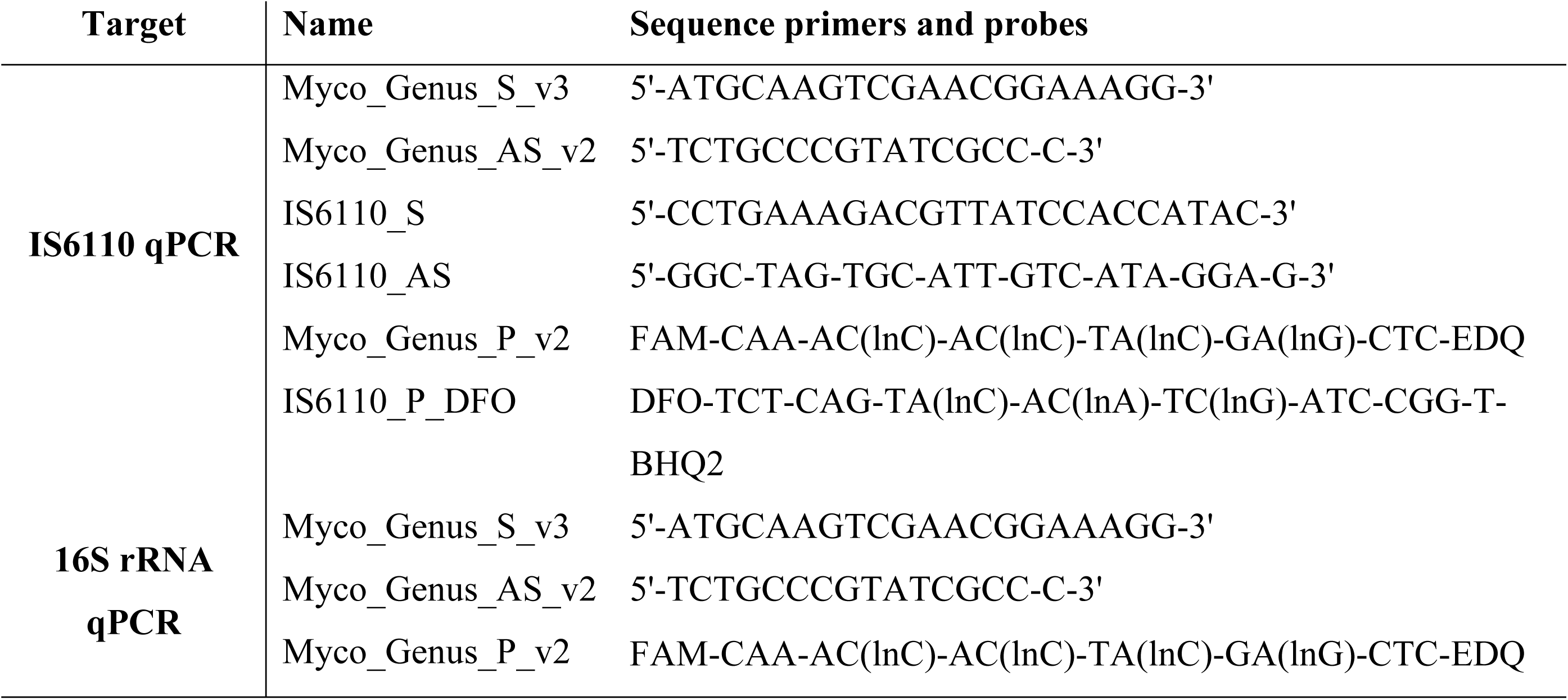
Primers and probes for qPCR to simultaneously detect *Mycobacterium tuberculosis* complex and all mycobacterial species.

**Table 2.** Interpretation of duplex IS*6110*-16SrRNA qPCR results.

| Cycle threshold (Ct) 16S rRNA | qPCR IS6110 result |  |
| --- | --- | --- |
|  | Positive | Negative |
| **Positive with any Ct value** | - | No MTBc |
| **Positive and ≥IS6110 Ct** | MTBc |  |
| **Positive and $<$IS6110 Ct** | MTBc + NTM |  |
| **Negative** | MTBc<br>(low bacillary load) | No mycobacteria species<br>identification possible |

Subsequently, a nested conventional PCR targeting the 16S rRNA gene was performed on samples identified as NTM by the duplex qPCR. The primers and probes used were as follows:

- P1 (first PCR): 5’-TGCTTAACACATGCAAGTCG-3’
- P2 new (first PCR): 5’-TCTCTAGACGCGTCCTGTGC-3’
- P7 (nested PCR): 5’-CATGCAAGTCGAACGGAAAGG-3’
- P16 new (nested PCR): 5’-AAGCCGTGAGATTTCACGAACA3’

Amplicons that were clearly positive on the agarose gel underwent Sanger sequencing. A negative PCR result was interpreted as the absence of MTBc or NTM.

Sequencing was subcontracted to BaseClear (BE Leiden, The Netherlands). The results obtained after entering the sequence into the Basic Local Alignment Search Tool (BLAST), National Centre for Biotechnology Information (NCBI) were interpreted as follows:

- If a sequence match was found on the NCBI, the NTM species was identified.

○ In case of 100% similarity (no mismatches), the species name was assigned.
○ In case 1-3 bp mismatches with the closest species, the “species-like” name was assigned
- If no match or >3 bp difference with the closest species was found on NCBI, the result was interpreted as *Mycobacterium* species.
- If there were overlapping peaks in the sequence, indicating the presence of several NTMs, the result was interpreted as “Mixture of NTMs”.

### 2.5. Data analysis

The data collected using the survey forms and sample submission forms were entered into EpiInfo version 7.2.5.0. The variables analysed were age, sex, region of residence, history of illness and history of anti-TB treatment. Percentages, medians and means were calculated by crosstab, and p-values and odds ratios with 95% confidence intervals were generated using regression analysis in IBM SPSS Statistics version 21X64 (IBM Corp., NY, USA).

### 2.6. Ethical considerations

The drug-resistance survey protocol in which a secondary analysis of samples is described in the informed consent was accepted by the ethics committee of the National Institute of Public Health Research by decision No. 08/2018/CE-INRSP on March 21, 2018. The objectives and results of the study were clearly explained in the language understood by the patient, and consent was obtained from patients over 18 years of age. For patients under the age of 18, after obtaining the prior consent of the parent or companion, the child’s assent to participate in the study was obtained in the presence of a witness other than the parent or companion.

## 3. Results

### 3.1. Mycobacteria detected

A total of 1,418 sputum AFB-positive patients were enrolled in the drug-resistance survey, including 1,199 new cases, 211 previously treated cases and 8 whose previous treatment history was unknown (Figure 1). Based on Xpert results, 1296 (91.4%) patients were positive for MTBc while 122 (8.6%) remained negative.

**Fig 1.**
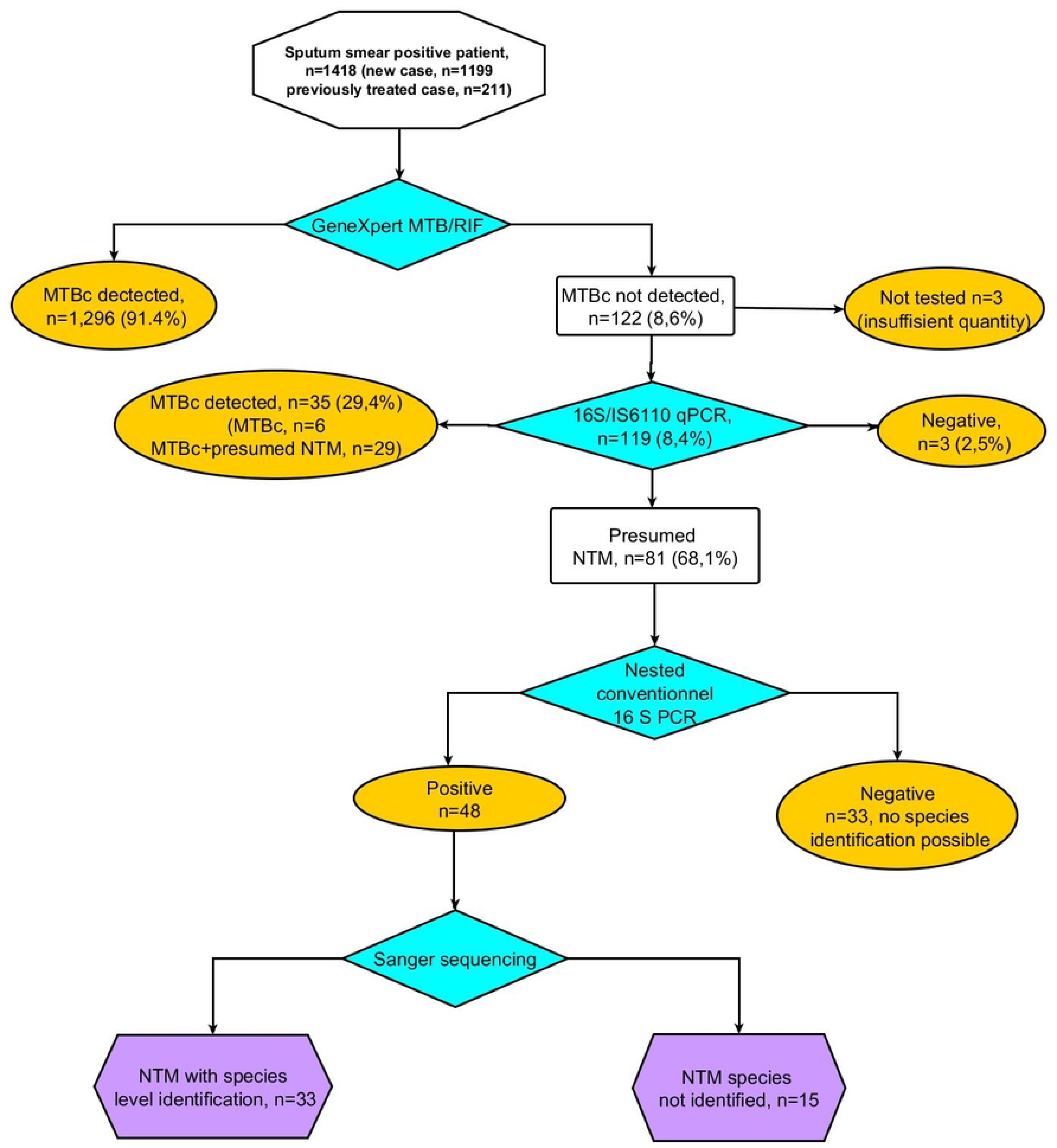
This is the Fig 1 **Sample and analysis flow with summary of results** This is the Fig 1 legend. *MTBc = Mycobacterium tuberculosis complex; n= number; NTM = non-tuberculous mycobacteria*.

The duplex IS*6110*-16S rRNA qPCR was performed for 119 patients with an Xpert result of “MTB not detected” by which increase the number of patients with MTBc increased to 1331 (93.9%) through 35 additional cases (29.4%), for six of whom MTBc alone was detected and 29 carrying probable mixtures of MTBc and NTMs. For 81 (68,1%) patients, only the 16S target reacted positive (presumed NTM), while the PCR remained negative for both targets in 3 (2,5%) patients. The nested conventional 16S PCR performed for the 81 presumed NTM detected by IS*6110*-16SrRNA qPCR, was positive for 48 or 3.4% (48/1418) of all enrolled patients, of which 33 could be identified to the species level by sequencing. For another 33 of 81 the nested PCR remained negative, thus no sequencing could be performed (Fig 1).

Sequencing identified eleven species or groups of species, with predominance of the *M. avium* complex (MAC) representing 20 cases or 42% of all species detected, *M. intracellulare* complex in 13 patients (27%), *M. avium* in five (10%) and *M. colombiense* in two (4 %) (Fig 2).

**Fig 2.**
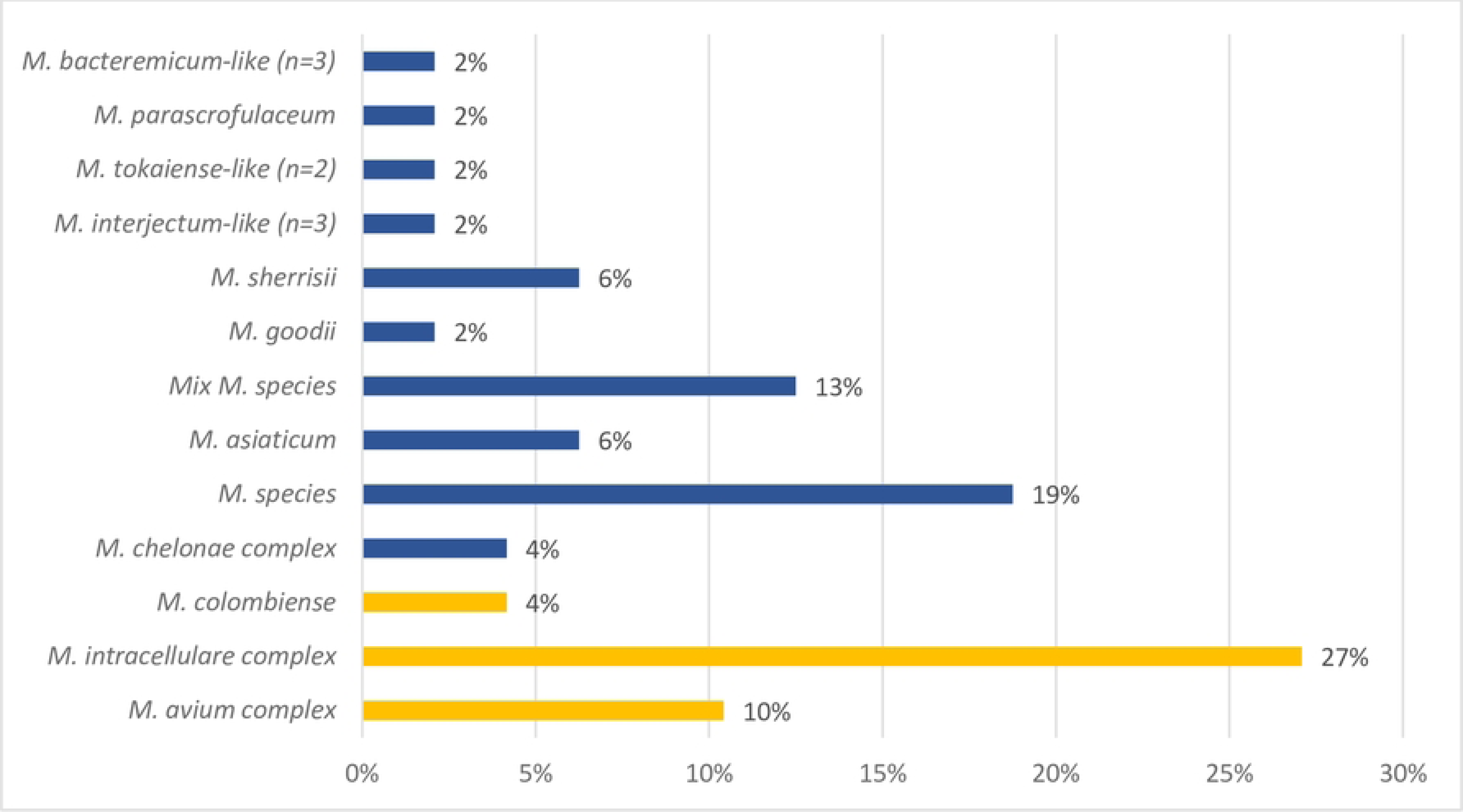
This is the Fig 2 **Sequencing results for the identification of NTM species**.

### 3.2. Detection of NTM in relation to smear-microscopy results

The quantification of AFB was notified for 1414 of 1418 patients by the participating laboratories. A significantly higher proportion of NTM was detected in patients with lower AFB-grades: 7% for 1+ and scanty (Table 3), compared to 2% among 3+ patients (respectively OR 4.065, 95% CI: 1.876-8.808, p=0.000 and OR 4.3, 95% CI: 2.012-9.189, p=0.000). The difference was not significant compared to grade 2+ samples (p=0.462).

**Table 3.** Proportion of NTM and MTB detection in relation to smear microscopy results.

| Final smear result | MTB n (%) | NTM n (%) | No species identification possible n (%) |
| --- | --- | --- | --- |
| **Rares BAAR** | 167 (83%) | 13 (7%) | 21 (10%) |
| **1+** | 170 (89%) | 14 (7%) | 8 (4%) |
| **2+** | 259 (94%) | 7 (3%) | 8 (3%) |
| **3+** | 731 (98%) | 14 (2%) | 2 (0%) |
| **Total** | **1327 (93.8%)** | **48 (3.4%)** | **39 (2.8%)** |

*MTBc = M. tuberculosis complex; NTM = non-tuberculous mycobacteria*

### 3.3. Socio-demographic, clinical characteristics and risk factors for NTM infection

Male patients were more numerous among all study participants with a sex ratio of respectively 2.1 and 1.5 when compared to women, but NTM presence was not associated with gender (Table 4). Contrary, older age was significantly associated with NTM detection. The median age was 37 years for all patients and 59.5 years for those infected with NTM detected. Compared to the under-34 age group, the 35-64 age group had a greater odds (OR 3.4, 95% CI: 1.1-10.3, p=0.03) of NTM detection, which further increased for the over-65 age group (OR 8.8, 95%CI: 2.3-33.2 p=0.001 (Table 4). Also, previously treated patients were more likely than new cases to have NTMs in their sputum (OR 3.4, 95%CI: 1.2-9.6, p=0.016) (Table 4). We found no significant associations between NTM detection and HIV status or chronic respiratory signs. Regarding the patient’s residence, patients from the Mopti region were more likely to have NTM detected compared to Bamako residents (OR 4.1, 95%CI: 1.0-16.4, p=0.04). (Table 4).

**Table 4.** Multivariate logistic regression analyses of demographic and clinical variables as risk factors of NTM infection.

| **Variables** |  | **Total**<br>**patients**<br>**n (%)** | **NTM**<br>**detected**<br>**n (%)** | **OR** | **p-value** |
| --- | --- | --- | --- | --- | --- |
| **Gender**<br>**n=1379** | Female | 434 (31.5) | 19 (39.6%) | 0.8 [0.3-2.04] | 0.7 |
|  | Male | 945 (68.5) | 29 (60.4%) |  |  |
| **Age n=1379** | Median | 37 | **59,5** |  |  |
|  | 0-34 years<br>(Reference) | 603 (43.7) | 4 (8.3) |  |  |
|  | 35-64 years | 659 (47.8) | 29 (60.4) | **3.4 [1.1-10.3]** | **0.03** |
|  | ≥65 years | 117 (8.5) | 15 (31.3) | **8.8 [2.3-33.2]** | **0.001** |
| **History of**<br>**treatment**<br>**n=1371** | New | 1165 (84.5) | 30 (62.5) |  |  |
|  | Previously<br>treatment | 206 (14.9) | 18 (37.5) | **3.4 [1.2-9.6]** | **0.016** |
| **HIV status**<br>**n=1092** | Positive | 77 (5.6) | 4 (8.3) | 2.8 [0.8-8.9] | 0.07 |
|  | Negative | 1015 (73.6) | 29 (60.4) |  |  |
| **Chronic**<br>**clinical signs**<br>**n=1174** | Yes | 442 (37.6) | 24 (60) | 0.7 [0.3-1.8] | 0.5 |
|  | No | 752 (62.4) | 16 (40) |  |  |
| **Region of**<br>**residence at**<br>**the time of**<br>**diagnosis**<br>**n=1348** | Bamako<br>(Reference) | 451 (33.5) | 5 (10.9) |  |  |
|  | Kayes | 101 (7.5) | 4 (8.7) | 2.7 [0.5-12.7] | 0.2 |
|  | Koulikoro | 217 (16.1) | 7 (15.2) | 1.5 [0.3-7.4] | 0.5 |
|  | Sikasso | 155 (11.5) | 12 (26.1) | 2.9 [0.7-10.8] | 0.10 |
|  | Ségou | 147 (10.9) | 7 (15.2) | 3.2 [0.8-12.7] | 0.09 |
|  | Mopti | 191 (14.2) | 7 (15.2) | **4.1 [1.02-16.4]** | **0.04** |
|  | Northern<br>regions | 86 (6.4) | 4 (8.7) | 4.2 [0.7-25.2] | 0.1 |

197 NTM = non-tuberculous mycobacteria; n= number; OR = odds ratio

## 4. Discussion

The rate of sputum AFB-positive patients and MTBc detected by Xpert in our study was slightly lower than the rate reported in other drug-resistance surveys, ranging from 95.8% −97.7%(14–17).

The use of an additional in-house qPCR for IS*6110* in our study has allowed to increase the detection of MTBc. The applied qPCR IS*6110* has shown good sensitivity and specificity for the detection of MTBc (18).The observed high Ct values indicates, however, a low quantity of MTB DNA anyway for samples from these patients, that may as well have become positive upon repeated Xpert testing(19).

The higher positivity rate by qPCR compared to the conventional nested PCR we observed in our study could be partially attributable to the non-specificity of the 16S primers employed for the in-house qPCR which also detects species belonging to closely related genera like *Rhodococcus* and *Nocardia*, hence potentially overestimating the NTM rate. These closely related species indeed can be detected in smear-positive samples not containing tubercle bacilli, such as documented in the USA with 78.4% (134/171) of AFB-positive specimens yielding a positive culture with NTM or *Nocardia* spp, while 14.6% (25/171) of AFB-positive specimens grew MTBc and another 7% (12/171) remained negative in culture (20). The proportion of NTM versus *Nocardia* spp note the clinical relevance of these isolated bacteria were discussed in that study. In another study in Nigeria, out of 32 smear-positive samples identified as not containing tuberculosis bacilli, 6 were NTM (18.75%), 14 were Corynebacteria (43.75%) and 12 Rhodococci (37.5%) (21). In our study, we did not further identify the non-MTBc/non-NTM samples. In addition, the lower sensitivity of the conventional nested PCR may have hampered confirmation by sequencing in case of negative results or low amplicon yield. Indeed, the in-house qPCR has already shown good sensitivity, while limited loss of DNA has been described during multiple handling steps for conventional nested PCR(22,23). This lower sensitivity is reflected in the samples’ AFB-grading, with less non-identified samples among the strong positive specimens.

The confirmed NTM detection rate in our study was higher (3.4%) than that obtained in TB drug resistance surveys conducted in Pakistan with 0.7% (13/1972) NTM (15,24), and in India with 1.19% (35/2,938) NTM (24), but it was lower than the 6.4% (317/4917) observed in China (25). This result may reflect regional differences in NTM occurrence and disease caused by NTMs around the world (26,27).

NTM detection was more likely among paucibacillary patients as determined by smear microscopy, corroborating findings from Nigeria where the AFB-smear grading was respectively lower for NTM cases than for MTBc patients (28) and was predictive for MTBc detection when it was higher (21)With age, the risk of developing NTMs infection increased, but we did not find a significant association between gender and NTM infection, contrary to other studies, where females have been more frequently associated with NTM infections, especially at older ages (6,29,30). Previous anti-TB treatment was another risk factor for NTM infection in our study, confirming findings from NTM pulmonary disease in China and South Africa (31,32). Nevertheless, we did not find an association between NTM detection and chronic respiratory signs in our study, in contrast with the described association between NTM infection and the presence of lung disease or chronic respiratory symptoms in Chinese and Danish studies(33,34).

HIV infection has frequently been identified as a risk factor for NTM infections and disease, especially in situations of advanced immunosuppression (28,35,36). In our study the trend towards an association between HIV-positivity and NTM detection was not significant, corroborating previous observation in Mali (37,38). In the context of the drug-resistance survey, no information on anti-retroviral treatment has been collected, but such treatment is to be started upon HIV diagnosis as per national guidelines in Mali. While patients from the Mopti region had a higher odds for having NTM detected in their sputum than those from the Bamako region, further investigation of the environmental conditions in these regions would be needed, especially as NTMs are environmental bacteria and have often been isolated at different frequencies in different geographical areas(26,30,39). The MAC was the most common NTM detected. The predominance of MAC has been reported in pulmonary NTM infections in many countries around the world (26,27,40,41) 29,38,40,41. In Mali, between 2004 and 2013, *M. avium* was the most frequently identified species of the MAC complex (7,27,37).

In this study, we did not investigate whether the detected NTM were attributable to true infection, colonization or sample contamination. We could not assess the AST criterium to detect NTM in at least 3 different samples per patient given the design of the survey and the retrospective nature of molecular analysis to detect NTM. However, study participants were people with respiratory signs undergoing evaluation for pulmonary TB attending health centers or patients failing TB treatment. Also, we do not describe the outcome of patients in whom NTM was detected, but all were started on 1st line TB therapy or multidrug-resistant treatment following the results of initial microscopy and Xpert testing.

## 5. Conclusion

Among sputum AFB-positive patients presenting in Mali, 3.4% were due to NTM. Particular attention must be paid to populations at risk, such as elderly patients, and patients with a history of previous treatment. TB prevention and care programmes in resource-limited countries should ensure that their routine diagnostic algorithms can detect such cases, based on the concurrent use of rapid molecular TB diagnostics and microscopy in the absence of other molecular assays to detect NTM. Clinical assessment and management of NTM infections however, remain a challenge worldwide.

## Data Availability

All data produced in the present study are available upon reasonable request to the authors

## Acknowledgements

*We would like to thank the tuberculosis management sites for recruiting patients, for collecting and transporting samples, the country’s regional health departments for regional coordination of the drug resistance study and Olga Tosas Auguet for her assistance in planning the survey, monitoring the field activities and preliminary data analysis”.*

